# Clinical Characterisation of Eleven Lateral Flow Assays for Detection of COVID-19 Antibodies in a Population

**DOI:** 10.1101/2020.08.18.20177204

**Authors:** Fabian Rudolf, Hans-Michael Kaltenbach, Janina Linnik, Marie-Therèse Ruf, Christoph Niederhauser, Beatrice Nickel, Daniel Gygax, Miodrag Savic

**Author notes:** these authors contributed equally (Fabian Rudolf, Hans-Michael Kaltenbach).

## Abstract

**Importance:** Serological assays can help diagnose and determine the rate of SARS-CoV-2 infections in a population.

**Objective:** We characterized and compared 11 different lateral flow assays for their performance in diagnostic or epidemiological settings.

**Design, Setting, Participants:** We used two cohorts to determine the specificity: (i) up to 350 blood donor samples from past influenza seasons and (ii) up to 110 samples which tested PCR negative for SARS-CoV-2 during the first wave of SARS-CoV-2 infections in Switzerland. The sensitivity was determined using up to 370 samples which tested PCR positive for SARS-CoV-2 during the same time and is representative for age distribution and severity.

**Main Outcome:** We found a single test usable for epidemiological studies in the current low-prevalence setting, all other tests showed lacking sensitivity or specificity for a usage in either epidemiological or diagnostic setting. However, orthogonal testing by combining two tests without common cross-reactivities makes testing in a low-prevalence setting feasible.

**Results:** Nine out of the eleven tests showed specificities below 99%, only five of eleven tests showed sensitivities comparable to established ELISAs, and only one fulfilled both criteria. Contrary to previous results from lab assays, five tests measured an IgM response in >80% of the samples. We found no common cross-reactivities, which allows orthogonal testing schemes for five tests of sufficient sensitivities.

**Conclusions and Relevance:** This study emphasizes the need for large and diverse negative cohorts when determining specificities, and for diverse and representative positive samples when determining sensitivities of lateral flow assays for SARS-CoV-2 infections. Failure to adhere to statistically relevant sample sizes or cohorts exclusively made up of hospitalised patients fails to accurately capture the performance of these assays in epidemiological settings. Our results allow a rational choice between tests for different use cases.

## Introduction

Antibodies are a hallmark of the human adaptive immune response to viral infections,^1^ and the immune system produces a heterogeneous population of different types of antibodies with distinct response kinetics and binding affinities.^2^ For instance, IgM is the first type of antibody induced upon infection, but has the shortest residence time and 2weakest antigen interaction. In contrast, IgA and IgG antibodies are produced later, but show orders of magnitude higher affinity and can provide long-term protection. Hence, IgM levels are sometimes used for diagnostic testing, while IgA and especially IgG levels can confirm a previous infection or vaccination.^3^

Scalable and accurate serological tests are fundamental to the understanding of the cumulative incidence of infections in a population.^4^ According to the WHO such knowledge can help in establishing the occurrence of infection in a population,^5^ which is crucial to determine the true extent of the disease, the distribution of severe, mild or asymptomatic cases, the infection fatality ratio of a population, the number of cases missed using routine disease surveillance methods, and the proportion of the population potentially protected against future infection.

A SARS-CoV-2 infection can lead to a strong IgG and IgA response for all tested epitopes in people with severe symptoms, while oligosymptomatic cases showed a diverse response in both antibody levels and epitope recognition.^6-8^ Most studies also point to a fast IgA and IgG response and a rather weak IgM response.

Lateral flow assays (LFA) are scalable and affordable tests^9^ that can potentially be used as both diagnostic and epidemiological assays. To be of use, those tests need to have a specificity >99.5% and a sensitivity >90%.^10^ The use and approval of a test by the FDA therefore requires a rigorous characterisation of its specificity, the probability that a negative sample yields a negative test result, and its sensitivity, the probability that a positive sample yields a positive test result. These characteristics also provide a means to compare different assays in the absence of established clinical decision points.

An accurate and precise determination of the specificity hinges on sufficiently many and sufficiently diverse samples to check for unknown cross-reactivities, such as a large, diverse panel of samples from blood donors including samples from flu seasons of the previous years, as performed for the Roche assay.^11^ Similarly but using a much smaller panel, post-market validations in Australia^12^ and the US^13^ are conducted by the regulators to check the products for their actual performance. Most LFA did not reach the manufacturers specification and were deemed “should not be used” in the US.^14^ Such cohorts also mimic the setting of a seroprevalence study, an important feature for epidemiological application of a test,^15^ where current low prevalence settings require high precision of specificity estimates.^16^

On the other hand, the sensitivity needs to be assayed using a sufficiently large and representative sample of the antibody response in a population for time post infection, (but especially also in disease severity) i don’t know what you mean. ^17^ A diverse standard panel representative for the SARS-CoV-2 response in a population yields a sufficiently precise estimate. However, most published clinical characterisations of LFA and claims from vendors are not based on such representative samples, but instead use small negative and/or positive cohorts predominantly comprised of hospitalised patients with severe cases of the disease.^18-22^ They thereby neglect the approximately 80% oligosymptomatic cases observed in the population^23^ and likely overestimate a the sensitivity of a test when applied population-wide rather than for severe cases only.

Here we present the clinical characterisation of eleven commercially available SARS-CoV-2 lateral flow assays (Table 1). All tests were characterised using the same positive cohort consisting of all or part of a collection of 366 convalescent samples^24^ and the same negative cohort of up to 500 blood donor samples from the influenza seasons 2016/17 and 2017/18. This allows a direct comparison of different characteristics of the tests.

**Table 1:** Lateral flow assays with vendor specifications. The manufacturer, buffer volume, volumes for serum(S), plasma (P), whole blood (WB), and incubation time are listed. Epitopes and production hosts are taken from the manufacturers documentation if available.

| Name | Buffer<br>μl (Drops) | S/P<br>μl | WB<br>μl | Time<br>min | Epitope | Host | SE<br>% | SP<br>% |
| --- | --- | --- | --- | --- | --- | --- | --- | --- |
| Lumiratek | 80 (2) | 10 | 10 | 10 | Spike |  | 93.53 | 97.50 |
| Sure Biotech | 80–100 (2) | 10 | 20 | 15–20 | S1, S2, RBD |  | 93.00 | 97.50 |
| Hightop | 80–100 (2) | 10 | 20 | 15–20 | Spike, NCP |  | 93.00 | 97.50 |
| Biozek | 80 (2) | 10 | 20 | 10 |  |  | 100.00 | 98.00 |
| Biotime | 100 (3) | 10 | 10 | 10 |  |  | 96.40 | 98.70 |
| TAmiRNA | 120 (3) | 10 | 20 | 10–15 | S1 | HEK-293 | 98.40 | 100.00 |
| NTBIO | 70–100 (2) | 10 | 10 | 15 |  |  | 97.83 | 98.00 |
| MEDSan | 60–80 (2) | 5 | 5 | 10–15 |  |  | 97.10 | 100.00 |
| MEXACARE | 90 (2) | 10 | 20 | 15–20 | RBD, NCP | *E.coli*, Eukaryotic cells | 97.65 | 99.53 |
| CTK Biotech | 70–100 (2) | 10 | 20 | 10–15 | Spike | Mouse | 96.00 | 97.80 |
| Augurix | 120 (3) | 10 | 20 | 10–15 | RBD, NCP | *E.coli*, 6his-Tag | 98.50 | 96.00 |

Overall, most tests appear unsuitable for general use: only two tests, Hightop and Augurix, achieved a specificity higher than 99%. Several tests, including Hightop, showed sensitivities greater than two previously characterized Euroimmun and Epitope Diagnostics ELISA (>92%), while Augurix showed a sensitivity only slightly above 50%. However, our analysis and data provide evidence that orthogonal testing strategies combining several tests with high sensitivity can compensate for the individual low specificities to achieve a combined specificity of more than 98.5% with sensitivities between 85-95% for IgG. Interestingly, we found that five LFA were able to detect an IgM response early after symptom onset in more than 80% of the samples. In contrast to previous findings, this indicates a robust IgM response in SARS-CoV-2 infections. Taken together, our study highlights the need for standardized testing of these assays and suggests applications for those that perform better performing.

## Material & methods

### Clinical specificity & sensitivity

The clinical specificity Sp=TN/(TN+FP) characterizes the qualitative performance of a dichotomous test based on the number of true negatives (TN) and false positives (FP) and is the probability that a sample with no or very low antibodies yields a negative test result.

Conversely, clinical sensitivity Sp=TP/(TP+FN) characterizes the probability that a patient with detectable level of antibodies yields a positive test result based on the number of true positives (TP) and false negatives (FN).

### Cohorts

To accurately calculate the specificity, we used the plasma of a *blood donor* cohort composed of donations from December 2016, February 2017, and February 2018. Additionally, we used the serum of our previously described positive (*SERO-BL-positive*) and negative (*SERO-BL-negative*) cohorts of study participants testing PCR-positive (resp. - negative) for SARS-CoV-2 during the initial wave of COVID-19 infections in the canton of Basel-Landschaft,^24^ Switzerland. We also recorded sample characteristics in regard to lipophilic appearance and hemolysis.

### Assay procedure

We characterised eleven different commercially available lateral flow assays (LFA) for detection of SARS-CoV-2 specific IgM and IgG with serum samples from the three cohorts (Table 1).

All LFA were performed according to their respective manual. In brief, test components were brought to room temperature, sera or plasma aliquots were completely thawed before testing. The test cassette was removed from the sealed pouch and the required amount of sample was pipetted into the specimen well (Table 1), followed by addition of two or three drops of sample buffer to the specimen or, if present, buffer well. Results were read within the specified time window stated in Table 1. Lot numbers and expiration dates are given in (Supp. Table 2)

Presence of bands was visually inspected, and each test was imaged with a digital camera (different models) under standardized lightning conditions. We considered a test valid if its control band was present, and we considered a valid test positive for the respective antibody if the SARS-CoV-2 specific IgM, IgG or IgM/IgG band was detected in the sample.

We assayed the Hightop test using whole blood,^24^ serum and plasma, while all other tests were assayed using serum and plasma. The Hightop and MEDSan assays were characterised at the SwissTPH using the identical biobank and experimental setup as outlined previously.^24^ Eight tests were characterised simultaneously at the KUSPO Miinchenstein and the Biotime at the FHNW, Muttenz. These latter nine tests were characterised using the experimental design outlined below.

### Experimental design

We prepared ten 96-well plates to distribute the samples of the SERO-BL-negative, SERO-BL-positive and blood donor cohorts. We aimed at a roughly equal distribution of IgG levels (as previously established using ELISA) on each plate to avoid plate-specific biases and to this end divided the positive samples into five strata of different IgG level, each strata occurring on each plate roughly the same number of times. Assignment of samples of each stratum and the negative cohorts to each plate was fully randomized. We selected two samples from the SERO-BL-negative cohort, and two samples each from medium and high-level IgG patients and replicated each of these samples on each plate to estimate between-plate variation. We also selected one patient sample at random for each plate and replicated it five times on that plate to provide an estimate of within-plate variation. Finally, we randomly selected one patient sample with high IgG level for each plate, and added a ten-fold 1:2 dilution series on the same plate to be able to establish detection limits for each test. Assignment of patient samples to wells was fully randomized individually for each plate and the same plate layouts were used for all tests.

### Statistical analysis

Data analysis and creation of figures and tables was carried out using in-house scripts in R;^25^ binomial confidence intervals are 95%-Clopper-Pearson intervals calculated using **exactci** provided by the package PropCIs.^26^

### Data and code availability

Data, results, and R scripts are available on GitLab: https://gitlab.com/csb.ethz/serobl-covid19-lta.

## Results

### Specificity

We calculated the specificities for all 11 LFA separately for the IgM and IgG responses (Table 2 and Figure 1). Note that the TAmiRNA assay uses a combined IgM/IgG response using a single band and values for IgM and IgG therefore coincide for this test.

**Table 2:** Overview on specificities. Number of true negatives (TN), false positives (FP), and resulting specificity Sp with 95%-confidence interval for each test for IgM and IgG based on all negative samples (left) and samples from blood donors only (right). Note that TAmiRNA uses a combined IgM-IgG readout, resulting in identical values for TN/FP/Sp.

| POCT | All negative cohorts |  |  |  |  |  | Blood donors from flu seasons 2016-2018 |  |  |  |  |  |
| --- | --- | --- | --- | --- | --- | --- | --- | --- | --- | --- | --- | --- |
|  | IgM |  |  | IgG |  |  | IgM |  |  | IgG |  |  |
|  | TN | FP | Sp [CI], % | TN | FP | Sp [CI], % | TN | FP | Sp [CI], % | TN | FP | Sp [CI], % |
| Lumiratek | 210 | 18 | 92.1 [87.8, 95.3] | 220 | 8 | 96.5 [93.2, 98.5] | 152 | 11 | 93.3 [88.2, 96.6] | 160 | 3 | 98.2 [94.7, 99.6] |
| Sure Biotech | 375 | 5 | 98.7 [97.0, 99.6] | 369 | 11 | 97.1 [94.9, 98.5] | 268 | 2 | 99.3 [97.3, 99.9] | 262 | 8 | 97.0 [94.2, 98.7] |
| Hightop | 261 | 0 | 100.0 [98.6, 100.0] | 260 | 1 | 99.6 [97.9, 100.0] | 150 | 0 | 100.0 [97.6, 100.0] | 149 | 1 | 99.3 [96.3, 100.0] |
| Biozek | 184 | 6 | 96.8 [93.3, 98.8] | 176 | 14 | 92.6 [87.9, 95.9] | 131 | 4 | 97.0 [92.6, 99.2] | 125 | 10 | 92.6 [86.8, 96.4] |
| Biotime | 274 | 8 | 97.2 [94.5, 98.8] | 269 | 13 | 95.4 [92.2, 97.5] | 180 | 6 | 96.8 [93.1, 98.8] | 178 | 8 | 95.7 [91.7, 98.1] |
| TAmiRNA | 363 | 8 | 97.8 [95.8, 99.1] | 363 | 8 | 97.8 [95.8, 99.1] | 260 | 5 | 98.1 [95.7, 99.4] | 260 | 5 | 98.1 [95.7, 99.4] |
| NTBIO | 355 | 21 | 94.4 [91.6, 96.5] | 336 | 39 | 89.6 [86.1, 92.5] | 253 | 15 | 94.4 [90.9, 96.8] | 238 | 29 | 89.1 [84.8, 92.6] |
| MEDsan | 245 | 15 | 94.2 [90.7, 96.7] | 243 | 17 | 93.5 [89.7, 96.1] | 144 | 6 | 96.0 [91.5, 98.5] | 142 | 8 | 94.7 [89.8, 97.7] |
| Mexacare | 325 | 30 | 91.5 [88.2, 94.2] | 349 | 6 | 98.3 [96.4, 99.4] | 225 | 21 | 91.5 [87.2, 94.6] | 241 | 5 | 98.0 [95.3, 99.3] |
| CTK Biotech | 344 | 9 | 97.5 [95.2, 98.8] | 346 | 7 | 98.0 [96.0, 99.2] | 245 | 6 | 97.6 [94.9, 99.1] | 246 | 5 | 98.0 [95.4, 99.4] |
| Augurix | 372 | 1 | 99.7 [98.5, 100.0] | 371 | 2 | 99.5 [98.1, 99.9] | 267 | 1 | 99.6 [97.9, 100.0] | 267 | 1 | 99.6 [97.9, 100.0] |

**Figure 1:**
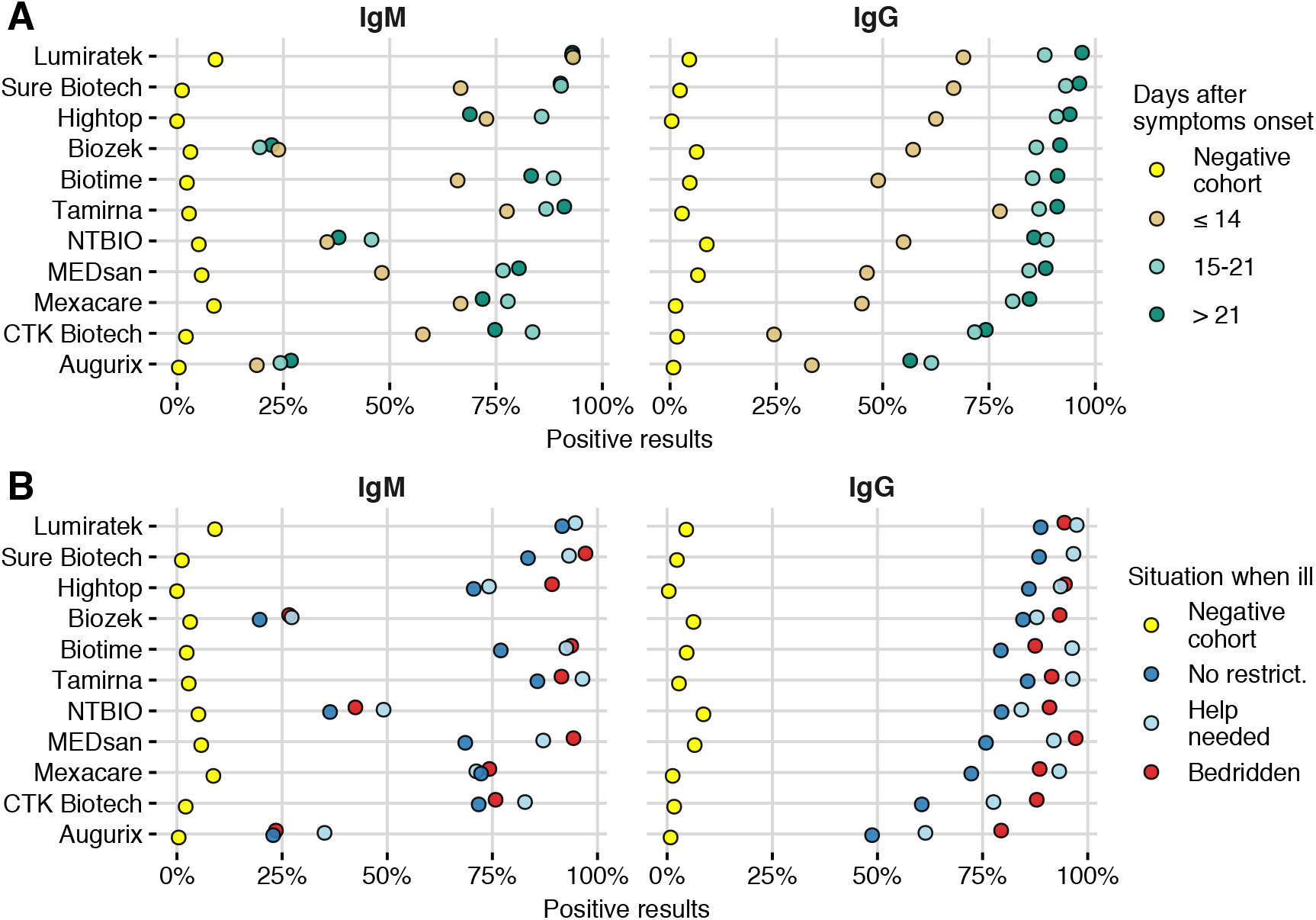
Overview on results. Specificities and sensitivities for all tests for IgM (left) and IgG (right). A: Sensitivities stratified by days after onset of symptoms. B: Sensitivities stratified by severity of disease.

Overall, the assays from Hightop and Augurix show specificities >99% for both IgG and IgM, while the assays by CTK, SureBiotech and Mexacare have specificities between 97% and 99% for IgG and IgM. The assay by Biotime also showed a specificity between 97% and 99%, but only for IgM. All other assays have specificities below 95% for both IgG and IgM. The combined IgG/IgM specificity of TAmiRNA is 97.8%.

To address potential cross-reactivity with different viruses circulating in the population, we additionally characterized each assay based only on blood donor samples from previous flu seasons. We found no significant differences for any test based on IgM, and only Lumiratek and MEDsan showed markedly worse characteristics for samples from earlier flu seasons.

We next checked whether samples are cross-reactive in multiple tests, which could be indicative for common epitopes, membranes or impurities. We found no noticeable shared cross-reactivities: Only 11 samples were positive in two or more tests and only two tests showed common cross-reactive samples for IgG (Supp. Table 5). Similarly, only six samples were positive in two or more IgM tests and only two tests showed common cross-reactive samples for IgM (Supp. Table 6). Only two samples were cross-reactive in two or more tests for both IgM and IgG.

### Sensitivity

The sensitivity of an assay depends on the time post infection and on the disease severity. To arrive at a comprehensive characterization of the sensitivity of each assay, we used samples from our previously established biobank of the canton of Basel-Landschaft. It contains samples from people tested during the first wave of the pandemic in Switzerland, with a wide range of days post symptom onset and of disease severity; these samples are representative for symptomatic and oligosymptomatic cases.^24^ Days post symptoms and disease severity were established with a doctor’s interview, and we categorized the severity in ‘bedridden’, ‘help needed’, and ‘no restriction’. An overview of the results stratified by days after onset of symptoms (Figure 1A) and severity of the disease (Figure 1B).

A positive IgG assay is considered as indicative of a past infection, and IgG seroconversion was previously reported measurable >14d post symptom onset and completed >21d (Figure 2, Supp Table 3). We therefore estimated the sensitivity of each test for IgG based on samples with more than 21 days post symptom onset (Table 3).

**Figure 2:**
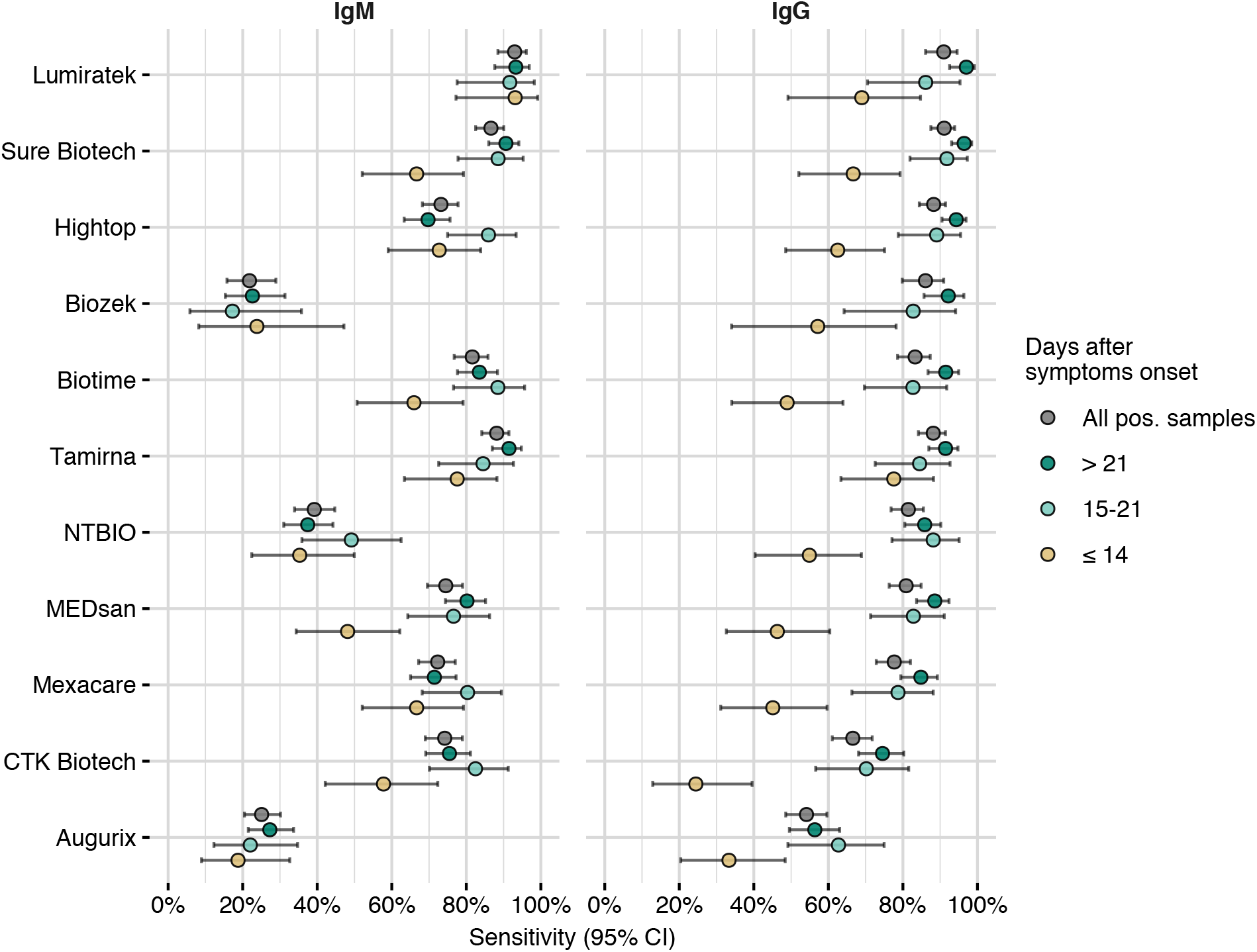
Sensitivity of LFA. The LFA results of all samples from the SERO-BL-positive cohort displayed by days post symptom onset for both IgG and IgM. TAmiRNA is identical in both panels, as it detects the combination of the two antibody type

**Table 3:** Sensitivity for IgG and >21 days post symptoms. Results for all positive samples and samples stratified by disease severity.

|  | All | Bedridden | Help needed | No restrictions |
| --- | --- | --- | --- | --- |
| **Lumiratek** |  |  |  |  |
| TP | 130 | 14 | 26 | 90 |
| FN | 4 | 0 | 0 | 4 |
| Se [CI], % | 97 [92.5, 99.2] | 100 [76.8, 100] | 100 [86.8, 100] | 95.7 [89.5, 98.8] |
| **Sure Biotech** |  |  |  |  |
| TP | 216 | 26 | 42 | 148 |
| FN | 8 | 0 | 1 | 7 |
| Se [CI], % | 96.4 [93.1, 98.4] | 100 [86.8, 100] | 97.7 [87.7, 99.9] | 95.5 [90.9, 98.2] |
| **Hightop** |  |  |  |  |
| TP | 216 | 26 | 41 | 149 |
| FN | 13 | 1 | 2 | 10 |
| Se [CI], % | 94.3 [90.5, 96.9] | 96.3 [81, 99.9] | 95.3 [84.2, 99.4] | 93.7 [88.7, 96.9] |
| **Biozek** |  |  |  |  |
| TP | 106 | 13 | 21 | 72 |
| FN | 9 | 0 | 3 | 6 |
| Se [CI], % | 92.2 [85.7, 96.4] | 100 [75.3, 100] | 87.5 [67.6, 97.3] | 92.3 [84, 97.1] |
| **Biotime** |  |  |  |  |
| TP | 183 | 21 | 39 | 123 |
| FN | 17 | 2 | 1 | 14 |
| Se [CI], % | 91.5 [86.7, 95] | 91.3 [72, 98.9] | 97.5 [86.8, 99.9] | 89.8 [83.4, 94.3] |
| **TAmiRNA** |  |  |  |  |
| TP | 203 | 24 | 40 | 139 |
| FN | 19 | 2 | 1 | 16 |
| Se [CI], % | 91.4 [87, 94.8] | 92.3 [74.9, 99.1] | 97.6 [87.1, 99.9] | 89.7 [83.8, 94] |
| **NTBIO** |  |  |  |  |
| TP | 188 | 22 | 33 | 133 |
| FN | 31 | 2 | 8 | 21 |
| Se [CI], % | 85.8 [80.5, 90.2] | 91.7 [73, 99] | 80.5 [65.1, 91.2] | 86.4 [79.9, 91.4] |
| **MEDsan** |  |  |  |  |
| TP | 201 | 26 | 40 | 135 |
| FN | 26 | 0 | 3 | 23 |
| Se [CI], % | 88.5 [83.7, 92.4] | 100 [86.8, 100] | 93 [80.9, 98.5] | 85.4 [79, 90.5] |
| **Mexacare** |  |  |  |  |
| TP | 190 | 24 | 40 | 126 |
| FN | 34 | 2 | 3 | 29 |
| Se [CI], % | 84.8 [79.4, 89.3] | 92.3 [74.9, 99.1] | 93 [80.9, 98.5] | 81.3 [74.2, 87.1] |
| **CTK Biotech** |  |  |  |  |
| TP | 158 | 22 | 33 | 103 |
| FN | 54 | 2 | 9 | 43 |
| Se [CI], % | 74.5 [68.1, 80.2] | 91.7 [73, 99] | 78.6 [63.2, 89.7] | 70.5 [62.4, 77.8] |
| **Augurix** |  |  |  |  |
| TP | 124 | 20 | 23 | 81 |
| FN | 96 | 6 | 19 | 71 |
| Se [CI], % | 56.4 [49.5, 63] | 76.9 [56.4, 91] | 54.8 [38.7, 70.2] | 53.3 [45, 61.4] |

The assays from SureBiotech, Lumiratek, and Hightop showed sensitivities >94%, a value exceeding the sensitivities of two previously characterised ELISAs (Epitope Diagnostics and Euroimmun),^24^ while the assay from Biotime showed a sensitivity of about 91%, comparable to these ELISAs. Tests generally showed higher sensitivities with increasing days post onset of symptoms, while Augurix and NTbio surprisingly detected more cases at >14d, although on a low level. The assays from SureBiotech and Lumiratek additionally showed a response for about 2/3 of the samples with less than 14 days post symptom onset.

Levels of IgM are expected to increase early during a disease and then decrease at later timepoints. We also observe this general trend for the 11 tests, even though each test seems to react differently at different stages (Supp Table 3). Detection of IgM levels is most important early in the disease. Between 7 and 28 days post onset of symptoms, only the assay by Lumiratek showed a sensitivity >90%, while tests from Sure Biotech, Hightop and Biotime still showed >80% sensitivity (Table 4). Considering the whole range of time points, the Lumiratek assay consistently showed sensitivities of about 90%, and tests by CTK, SureBiotech, Mexacare, Biotime, and Hightop had sensitivity >83% in at least one of the time windows. The combined detection of IgM and IgG for TAmiRNA results in a monotone increase of the sensitivity with time, starting from about 78%.

**Table 4:** Sensitivity for IgM and 7-28 days post symptoms. Results for all positive samples and samples stratified by disease severity.

|  | All | Bedridden | Help needed | No restrictions |
| --- | --- | --- | --- | --- |
| **Lumiratek** |  |  |  |  |
| TP | 113 | 8 | 21 | 84 |
| FN | 10 | 0 | 0 | 10 |
| Se [CI], % | 91.9 [85.6, 96] | 100 [63.1, 100] | 100 [83.9, 100] | 89.4 [81.3, 94.8] |
| **Sure Biotech** |  |  |  |  |
| TP | 176 | 19 | 31 | 126 |
| FN | 30 | 0 | 1 | 29 |
| Se [CI], % | 85.4 [79.9, 90] | 100 [82.4, 100] | 96.9 [83.8, 99.9] | 81.3 [74.2, 87.1] |
| **Hightop** |  |  |  |  |
| TP | 173 | 20 | 29 | 124 |
| FN | 43 | 0 | 6 | 37 |
| Se [CI], % | 80.1 [74.1, 85.2] | 100 [83.2, 100] | 82.9 [66.4, 93.4] | 77 [69.7, 83.3] |
| **Biozek** |  |  |  |  |
| TP | 19 | 2 | 4 | 13 |
| FN | 80 | 4 | 14 | 62 |
| Se [CI], % | 19.2 [12, 28.3] | 33.3 [4.3, 77.7] | 22.2 [6.4, 47.6] | 17.3 [9.6, 27.8] |
| **Biotime** |  |  |  |  |
| TP | 150 | 15 | 26 | 109 |
| FN | 31 | 1 | 2 | 28 |
| Se [CI], % | 82.9 [76.6, 88.1] | 93.8 [69.8, 99.8] | 92.9 [76.5, 99.1] | 79.6 [71.8, 86] |
| **TAmiRNA** |  |  |  |  |
| TP | 172 | 17 | 27 | 128 |
| FN | 28 | 2 | 2 | 24 |
| Se [CI], % | 86 [80.4, 90.5] | 89.5 [66.9, 98.7] | 93.1 [77.2, 99.2] | 84.2 [77.4, 89.6] |
| **NTBIO** |  |  |  |  |
| TP | 88 | 11 | 17 | 60 |
| FN | 114 | 7 | 14 | 93 |
| Se [CI], % | 43.6 [36.6, 50.7] | 61.1 [35.7, 82.7] | 54.8 [36, 72.7] | 39.2 [31.4, 47.4] |
| **MEDsan** |  |  |  |  |
| TP | 155 | 19 | 31 | 105 |
| FN | 60 | 0 | 4 | 56 |
| Se [CI], % | 72.1 [65.6, 78] | 100 [82.4, 100] | 88.6 [73.3, 96.8] | 65.2 [57.3, 72.5] |
| **Mexacare** |  |  |  |  |
| TP | 156 | 15 | 24 | 117 |
| FN | 50 | 4 | 8 | 38 |
| Se [CI], % | 75.7 [69.3, 81.4] | 78.9 [54.4, 93.9] | 75 [56.6, 88.5] | 75.5 [67.9, 82] |
| **CTK Biotech** |  |  |  |  |
| TP | 151 | 14 | 29 | 108 |
| FN | 39 | 3 | 2 | 34 |
| Se [CI], % | 79.5 [73, 85] | 82.4 [56.6, 96.2] | 93.5 [78.6, 99.2] | 76.1 [68.2, 82.8] |
| **Augurix** |  |  |  |  |
| TP | 50 | 3 | 11 | 36 |
| FN | 149 | 15 | 20 | 114 |
| Se [CI], % | 25.1 [19.3, 31.7] | 16.7 [3.6, 41.4] | 35.5 [19.2, 54.6] | 24 [17.4, 31.6] |

Levels of both IgG and IgM are also expected to increase with disease severity, leading to higher sensitivity for increased severity (Figure 2, Supp. Table 4). Sensitivities for IgG in the bedridden cohort >21d was >92% for all tests except Augurix. For IgM, all tests except Augurix and Biozek had sensitivities >90% for IgM in the bedridden cohort <21d. Sample size are low in these cases, precluding a more robust analysis. For oligosymptomatic cases—the majority for COVID-19—only five assay (SureBiotech, Biozek, Lumiratek, Biotime, Hightop) showed sensitivities above 90% for IgG. Moreover, only the Lumiratek assay showed a sensitivity above 90% for IgM in the ‘no restriction’ cohort at less than 21 days post symptoms.

### Predictive value and usage

Sensitivity and specificity describe the probabilities of a test correctly recognizing a positive or negative sample. For applications, especially serological studies, we are more interested in the converse conclusion: given a positive or negative test result, how likely is it that the underlying sample is truly positive or negative, respectively? These probabilities are given by the *positive predictive value (PPV)* and the *negative predictive value (NPV)*, respectively. Their values depend on the sensitivity and specificity of a test, but importantly also on the true prevalence of the disease in the population. They are calculated as

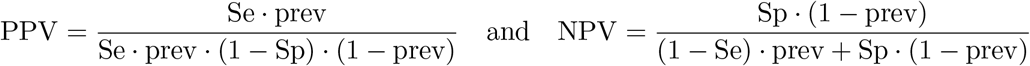

To evaluate the suitability of each test for applications, we calculated the PPV and NPV for an assumed prevalence between 0% and 25% (Figure 3 and Supp. Figure 8).

**Figure 3:**
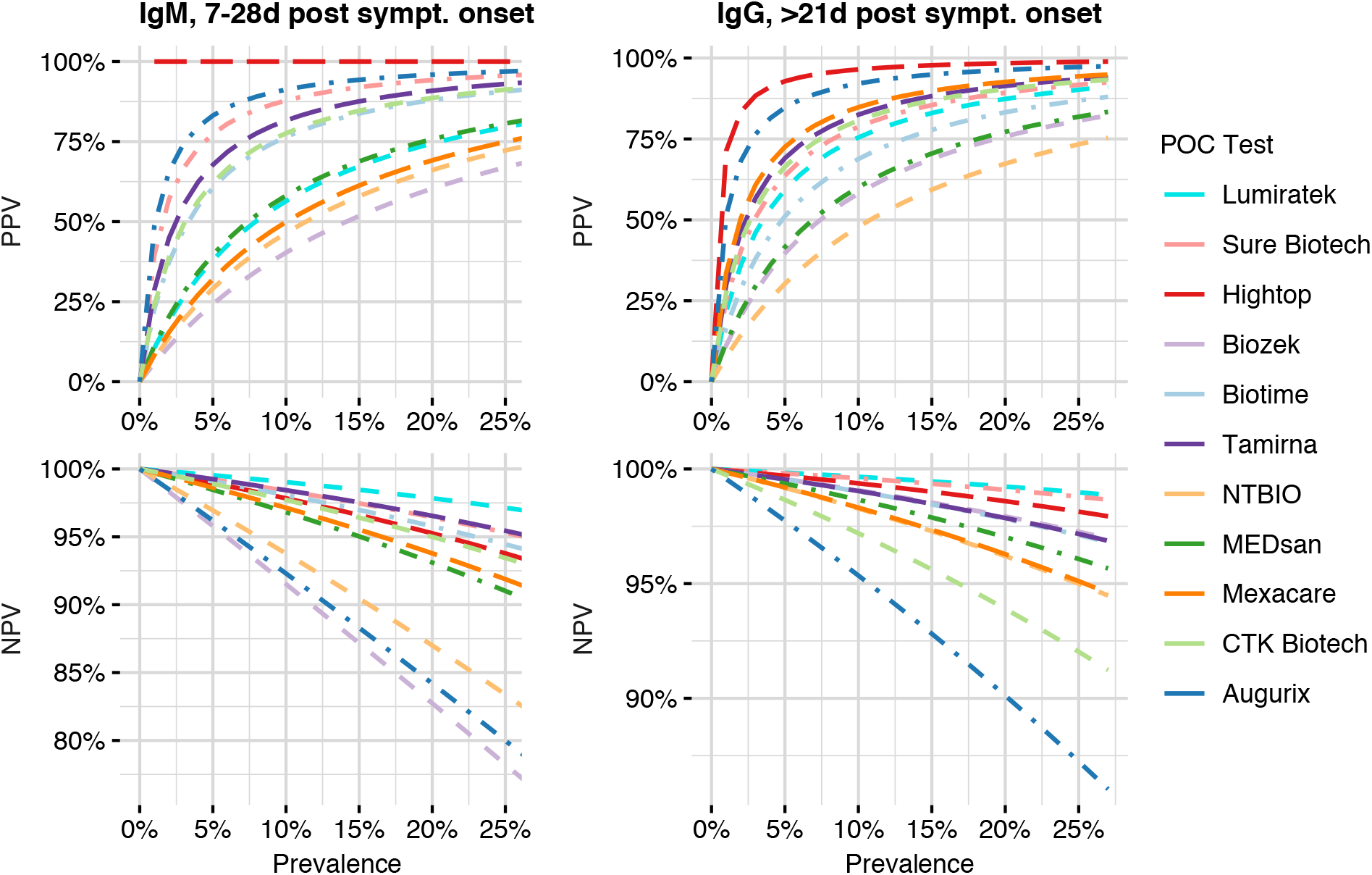
PPV and NPV for 0-25% prevalence. All negative samples have been included in the analysis. For IgM (IgG), only positive samples with 7-28 (>21) days after onset of symptoms were considered.

The PPV is mainly determined by the specificity of the test, and a low prevalence can severely influence the PPV due to the high proportion of false positive test results (Figure 3).

For IgG at >21d post onset of symptoms, only the assays by Hightop and Augurix showed sufficient PPV for low prevalence, while all other assays showed poor performance, often far below 50% PPV (meaning at least one in two positive tests is incorrect). Even at prevalence as high as 5%, the tests by MEDSan, Biotime, Biozek, and NTBio have PPVs below 50%. Results for IgM for 7-28 days post symptoms are generally worse.

Conversely, the NPV is largely determined by the sensitivity of a test and usually decreases with increasing prevalence, as proportionally more positive cases are observed; this decrease in NPV is therefore more pronounced for tests with low sensitivity, especially Augurix and CTK (Figure 3).

The PPV can be substantially increased by a strategy of *orthogonal testing*, where results of two or more tests are combined.^10^ For maximum effect, this strategy requires combining tests of reasonable sensitivity without shared cross-reactivities. We therefore selected the six individual tests with >92% sensitivity for IgG at >21 days post symptoms, respectively the four tests with IgM sensitivity >82% for 7-28 days post symptoms. The respective orthogonal tests based on pairs of tests almost all showed specificities >99.3%, the exceptions are Lumiratek combined with either SureBiotech or Biotime, which both showed specificities of ~98.5%. Naturally, the sensitivities decreased compared to individual tests, but remained >90% for some combinations. Overall, the specificities of the orthogonal tests are similar or better than the most specific single test—Hightop with 99.6% specificity. For IgM, we found combined specificities of around 99% with sensitivities of around 80%, still yielding workable PPVs (Table 5).

**Table 5:** Combining tests increases specificity. The tests with a sensitivity for IgG >92% and the IgM >82% were analysed for their combined specificty and sensitivity

| Test 1 | Test 2 | IgG |  |  |  |  |  | IgM |  |  |  |  |  |
| --- | --- | --- | --- | --- | --- | --- | --- | --- | --- | --- | --- | --- | --- |
|  |  | TN | FP | Sp [CI], % | TP | FN | Se [CI], % | TN | FP | Sp [CI], % | TP | FN | Se [CI], % |
| Lumiratek | Sure Biotech | 223 | 3 | 98.7 [96.2, 99.7] | 127 | 7 | 94.8 [89.5, 97.9] | 223 | 3 | 98.7 [96.2, 99.7] | 103 | 20 | 83.7 [76, 89.8] |
|  | Hightop | 154 | 1 | 99.5 [96.5, 100] | 125 | 9 | 93.3 [87.6, 96.9] |  |  |  |  |  |  |
|  | Biozek | 188 | 0 | 100.0 [98.1, 100] | 102 | 12 | 89.5 [82.3, 94.4] |  |  |  |  |  |  |
|  | Biotime | 166 | 2 | 98.8 [95.8, 99.9] | 102 | 8 | 92.7 [86.2, 96.8] | 166 | 2 | 98.8 [95.8, 99.9] | 79 | 19 | 80.6 [71.4, 87.9] |
|  | TAmiRNA | 219 | 1 | 99.5 [97.5, 100] | 119 | 13 | 90.2 [83.7, 94.7] | 218 | 2 | 99.1 [96.8, 99.9] | 95 | 22 | 81.2 [72.9, 87.8] |
| Sure Biotech | Hightop | 257 | 1 | 99.6 [97.9, 100] | 210 | 14 | 93.8 [89.7, 96.5] |  |  |  |  |  |  |
|  | Biozek | 188 | 1 | 99.5 [97.1, 100] | 104 | 11 | 90.4 [83.5, 95.1] |  |  |  |  |  |  |
|  | Biotime | 276 | 2 | 99.3 [97.4, 100] | 179 | 21 | 89.5 [84.4, 93.4] | 276 | 2 | 99.3 [97.4, 99.9] | 143 | 38 | 79.0 [72.3, 84.7] |
|  | TAmiRNA | 367 | 1 | 99.7 [98.5, 100] | 199 | 23 | 89.6 [84.7, 93.3] | 366 | 2 | 99.5 [98.1, 99.9] | 158 | 42 | 79.0 [72.7, 84.4] |
| Hightop | Biozek | 128 | 0 | 100 [97.2, 100] | 105 | 10 | 91.3 [84.6, 95.8] |  |  |  |  |  |  |
|  | Biotime | 172 | 1 | 99.4 [96.9, 100] | 175 | 25 | 87.5 [82.1, 91.7] |  |  |  |  |  |  |
|  | TAmiRNA | 250 | 0 | 100.0 [98.5, 100] | 194 | 28 | 87.4 [82.3, 91.5] |  |  |  |  |  |  |
| Biozek | Biotime | 143 | 0 | 100.0 [97.5, 100] | 77 | 14 | 84.6 [75.5, 91.3] |  |  |  |  |  |  |
|  | TAmiRNA | 185 | 0 | 100.0 [98, 100] | 96 | 17 | 85.0 [77, 91] |  |  |  |  |  |  |

## Discussion

Accurate and precise estimation of seroprevalence in a population is crucial for helping public health officials make informed decisions for targeting affected areas. Suitable serological tests have to accurately capture the spectra of the human antibody response in a population. Here, we assessed the performance of 11 commercially available lateral flow assays. We used samples from a previously established biobank of symptomatic and oligosymptomatic patients representative for the disease spectrum observed in western Europe, augmented by samples from blood donors of previous influenza seasons.

Our point estimates of specificities differ from those previously reported in studies or by manufacturers. However, the corresponding interval estimates show compatible estimates with all but one previous study (Supp Table 7). Our comparatively large number of negative samples resulted in a narrower interval estimation, indicating higher precision.

We did not observe clusters of samples showing cross-reactivity, these false positive samples are unlikely to come from a common recognition of the employed epitopes. This also indicates that few common cross-reactivities exist and specificities are therefore inherent to each lateral flow assay. Consequently, orthogonal testing strategies become viable options for increasing specificity beyond a single test. Our results indicate that specificity increases dramatically while the sensitivity remains usable, most often by employing a combination of the Spike and NCP as epitopes.

Our point estimates of the sensitivities differ substantially from previous reports for some tests (Supp Table 7), but interval estimates are again compatible. In particular, our estimated sensitivities of samples of bedridden patients with >21 days post symptoms are comparable to most previous studies. This subcohort includes hospitalised patients and might therefore be comparable to the previous studies, but no definite statement is possible due to the small size of this subcohort.

Importantly, our biobank allows stratifying the sensitivity estimates by days post symptoms and disease severity, thereby providing a more detailed picture of test performance for oligosymptomatic patients which comprise about 80% of cases in a population. We found that for the ‘no restriction’ cohort, three assays-SureBiotech, Lumiratek, Hightop-perform similarly to previously characterised ELISAs.

In contrast to previous reports, we were able to determine an early IgM response with several assays. This hints at the possibility of using IgM for diagnostic purposes in early infection, but our cohort is again for a conclusive statement. Additionally, the sensitive IgM assays show low specificity resulting in a high false positive rate, and their results can therefore only serve as an additional input for a clinical decision. At the moment, there is no diagnostic value in the IgG measurement as a comparison to neutralising titers is missing in all studies as well as the vendors specifications.

We anticipate that our characterisation of 11 LFA on common and diverse samples provides a basis to establish clinical and epidemiological decision points from which analytical sensitivities can be established. This requires standard panels of antibodies representative for the immune response to a SARS-CoV-2 infection, which are still under development.^10^ In contrast, the clinical specificity will always remain the sole meaningful one and future specificity tests have to be performed on rather large, diverse panels of negative samples, especially as only few common cross-reactivities were detected.

## Data Availability

Data are available upon request

## Acknowledgment

We thank Christian Kahlert for careful reading of the manuscript. The Swiss Red Cross financed all the used LFA except for the Hightop and Biotime assays. The Hightop was purchased by the canton Basel-Landschaft and the Biotime was provided by the Swiss importer. Thomas Büeler organised all the purchases, and helped in organising the logistics for the testing at the KUSPO Münchenstein site. Biolytix AG, Witterswil, Switzerland rearranged the whole biobank according to the experimental design used at the testing site. FR is funded by the NCCR ‘Molecular Systems Engineering’.

